# Variation of the vaginal microbiome during and after pregnancy in Chinese women

**DOI:** 10.1101/2020.07.07.20148536

**Authors:** Xiaoai Zhang, Qingzhi Zhai, Jinfeng Wang, Xiuling Ma, Bo Xing, Hang Fan, Zhiying Gao, Fangqing Zhao, Wei Liu

## Abstract

A more complete profiling of vaginal microbial communities and their variability enables a more accurate description of women microbiome. However, there is a distinct lack of information regarding the Chinese women. Composition of the vaginal microbiota during pregnancy and 6 weeks postpartum of 454 Chinese women thus was characterized in this study by sequencing V3-V4 regions of the 16S ribosomal RNA (rRNA). It showed that the vaginal microbiome varied during pregnancy and postpartum in response to abortion history, hypertensive disorders (HBP), delivery mode and maternal age. Co-variation of 21 bacterial taxa, including *Lactobacillus* and two of its species, may account for the common characteristics of vaginal microbiome under different medical histories and pregnancy outcomes. On the contrary, discriminant bacteria were significantly different between premature rupture of membranes related preterm birth (PROM-PTB) and non-PROM related PTB, and community state type (CST) I without any predominant *Lactobacillus* species in microbiota was more prevalent during pregnancy in PROM-PTB, suggesting that specific bacteria could be considered to distinguish different types of PTB. Through adding the data from Chinese women, the study will enrich the knowledge of human microbiome and likewise contribute to a better understanding of the association between the vaginal microbiome and reproductive health.

## Introduction

The Human Microbiome Project (HMP) has released the human microbiome data derived from thousands of individuals [1]. As an important part of the HMP and one of the most prosperous community in human body, vaginal microbiome has received much attention [2, 3]. Over recent years, evidence is accumulating that vaginal microbiome is key to women’s health and a healthy pregnancy, and people gradually realize that the health status largely depends on this microbial community harboring more beneficial commensals or pathogens [4-9]. Most of the studies have introduced the compositions and shifts of the vaginal microbiome in women of African, European and American population [10, 11], however, there is a lack of knowledge about the vaginal microbiome in Chinese pregnant women.

In China, nearly half of the births were delivered by caesarean section in 2007-2008, and the rate was even close to 60% in some cities [12-14]. With start of two-child policy in China from 2015, healthcare providers are facing the greater challenges of either a higher cesarean section rate, or an advanced maternal age, or both. In addition, in China, more than 1 million preterm infants are born per year, second only to India [15-17]. All these serious situations in obstetrics and gynecology have become major public health concerns in China and around the world. Previous studies have shown that the composition of vaginal microbiome was associated with pregnancy, delivery mode, maternal age, and preterm [4-8]. A more complete characterization of vaginal microbiome and its variability with pregnancy, delivery mode, or specific characters of pregnant women will enable a more accurate diagnosis of women who truly possess an abnormal vaginal microbiome.

In recent years, our understanding of vaginal microbiome has broadened by using cultivation intendent high-throughput sequencing. Ravel et al. performed a 16S ribosomal RNA (rRNA) gene survey on vaginal samples from 396 North American women represented four ethnic groups [18]. They divided the vaginal microbiome into five community state types (CSTs) based on the species of dominant bacteria. CST I, CST II, CST III and CST V was found to be predominated by *Lactobacillus crispatus, Lactobacillus gasseri, Lactobacillus iners*, and *Lactobacillus jensenii*, respectively, and CST IV was defined as lacking *Lactobacillus spp*. and comprising a diverse set of strict and facultative anaerobes. Following longitudinal study in white, black, and Hispanic reproductive-age women over 16 weeks, it was suggested that CSTs was dynamic in some women, but relatively stable in the others [19]. Thereafter, CSTs were widely used in the studies of the association between the vaginal microbiome and reproductive health for its effective handle.

To date, some studies linking the vaginal microbiome to preterm birth (PTB) have yielded mixed and even discordant results [4-7]. Romero et al. found no association between vaginal microbiome and either premature rupture of membranes (PROM) related or non-PROM related PTB in a predominantly African American cohort [4]. One study reported that CST V were associated with clinically heterogeneous PTB in two predominantly Caucasian populations [5]. Lindsay et al detected a significant positive association between CST III and non-PROM related PTB in high-risk Caucasian, Asian, and Black pregnant women [6]. Recently, Digiulio et al. replicated their previously reported associations [5] between less *Lactobacillus* or more *Garderella* and clinically heterogeneous PTB in the low risk Stanford cohort [7]. However, their previously hypothesized association [5] between PTB and more *Ureaplasma* was not replicated in their following study [7]. Resolving these mixed and even discordant findings in prior studies requires the need for investigation of the microbiome with different types of PTB and PROM simultaneously in a more comprehensive design and a larger sample size of cohort.

In this study, by using sequencing of 16S rRNA gene amplicons, we characterized and compared the vaginal microbiome community of Chinese pregnant women according to multiple factors such as delivery mode, maternal age, abortion history, pre-pregnancy maternal weight status, and pregnancy complications during pregnancy and postpartum. Furthermore, the relationships between vaginal microbiome and the clinical features of adverse pregnancy outcomes, in particular PTB, were analyzed. We wanted to expand the current understanding for the vaginal microbiome in Chinese pregnant women and whether the microbial community shifts over time or under certain conditions.

## Results

From July to December 2016, totally 474 pregnant Chinese women attending the Department of Obstetrics at the 301 Hospital (Beijing) for regular check-ups were enrolled in this study. Of the 474 volunteers recruited, 20 pregnant women were excluded because of medical complications (from fetal or pregnant women) requiring induction of labor. Of 454 women included in final analyses, vaginal swabs of 142 pregnant women were sampled at early stage of pregnancy (≤ 18 gestational weeks), 207 were sampled at late stage of pregnancy (27 ≤ gestational weeks < 42), and 98 were sampled 6 weeks postpartum (Supplementary Table S1). Using 16S rRNA-based sequencing we obtained a total of 27,171,551 high-quality sequences from the vaginal samples of Chinese pregnant women, with median 59,912 and interquartile range 57,311-62,351 sequences per sample. Totally 6343 operational taxonomic units (OTUs) were generated with ≥ 97% sequence similarity.

### The vaginal microbial community shifts significantly from pregnancy to postpartum

Alpha diversity, as quantified by the Chao1 index, was significantly higher (*P* < 0.001) in the vaginal microbiome postpartum than pregnancy (Figure 1A). The other four diversity indices showed consistent results (Supplementary Figure S1A-D), indicating that the microbial richness may have increased dramatically after delivery. Similarly, significant differences were also found in β diversities based on the weighted UniFrac dissimilarities (ANOSIM R = 0.511, *P* = 0.001) between pregnancy and postpartum (Figure 1B and Supplementary Figure S2A-D). Moreover, vaginal samples from pregnancy clustered more closely.

**Figure 1.**
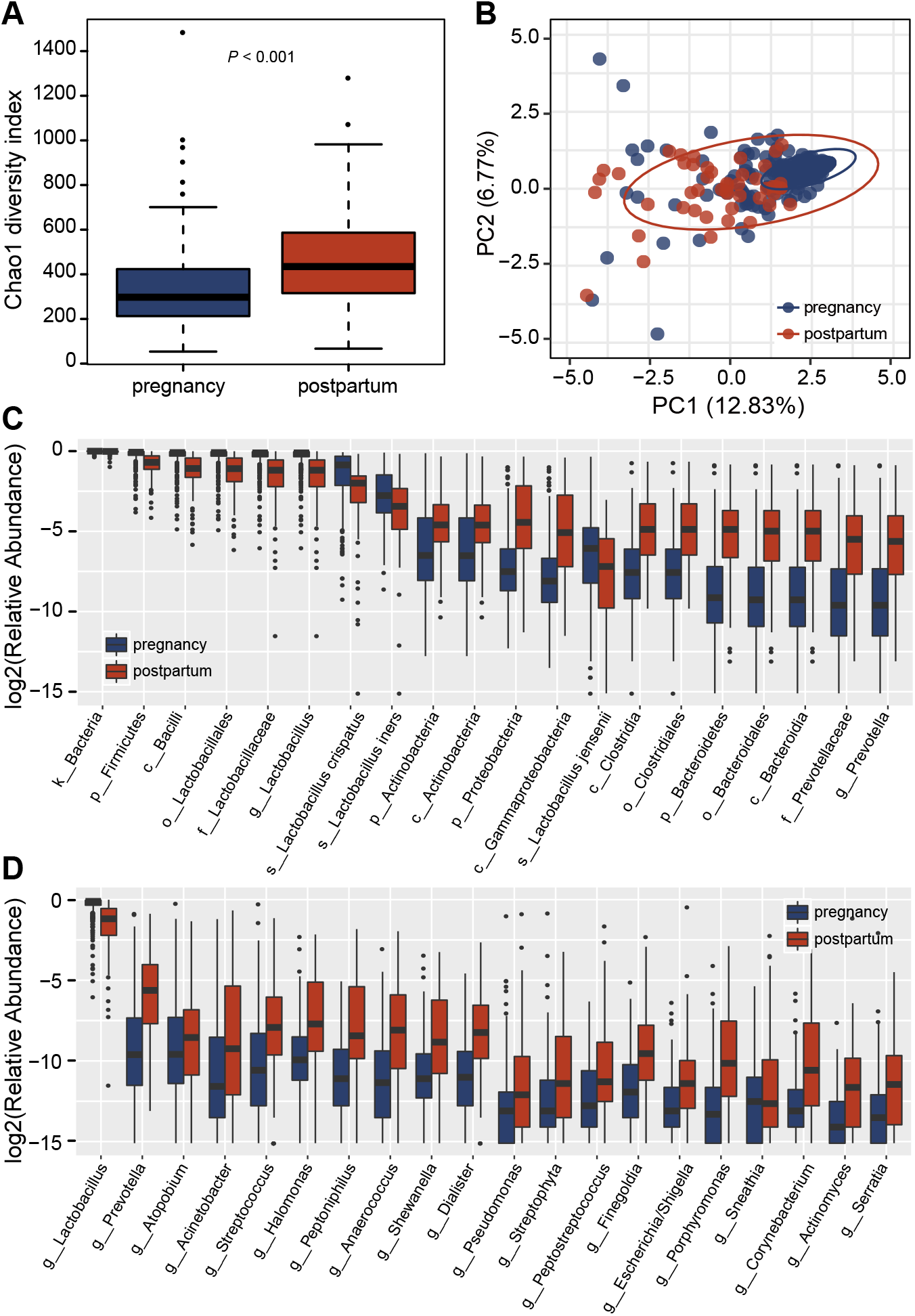
The vaginal microbiome during pregnancy and postpartum was significantly different in microbial diversity, community structure and composition. **(A)** Alpha diversity of the vaginal microbiome between pregnancy and postpartum. Each box plot represents the median, interquartile range, minimum, and maximum values. **(B)** Weighted ANOSIMs and principal coordinate analysis (PCoA) analysis of the vaginal microbiome during pregnancy and postpartum based on the distance matrix of UniFrac dissimilarity. The x- and y-axes represent two dimensions explaining the greatest proportion of variance in the communities. Each dot represents a sample, and each circle shows a 95% confidence interval. **(C)** Relative abundance of significantly different taxa during pregnancy and postpartum at all levels. **(D)** Relative abundance of significantly different taxa during pregnancy and postpartum at the genus level. The discriminating taxa were identified based on linear discriminant analysis effect size (LEfSe) analysis with a threshold of linear discriminant analysis (LDA) scores (log10) > 2 and *P* < 0.05. The prefixes p_, c_, o_, f_, g_, s_ represent phylum, class, order, family, genus, and species respectively.

For each taxonomic category, the difference was rather large. At the phylum level, only the relative abundance of Firmicutes was higher during pregnancy than postpartum, while more Actinobacteria, Proteobacteria, Bacteroidetes and other 7 phyla appeared in postpartum women (Figure 1C). *Lactobacillus* genus and 5 of its species *L. crispatus, L. gasseri, L. iner, L. jensenii*, and *L. reuteri* were significantly higher, and dozens of genera such as *Prevotella, Atopobium, Acinetobacter* and *Sneathia* were significantly lower during pregnancy (Figure 1D and Supplementary Table S2).

In view of the obvious difference of *Lactobacillus* before and after birth, we identified four CSTs from the vaginal microbiome of Chinese pregnant women (Supplementary Figure S3 and Supplementary Table S3). CST I and CST III were *L. crispatus* and *L. iners* dominated, respectively. Both CST IV-A and CST IV-B were non-*Lactobacillus* dominated, and the difference between them is that some women in the former still retained a certain amount of *L. crispatus, L. iners, L. jensenii* and/or *L. gasseri*, while the latter contained more *Gardnerella*. The most prevalent CSTs observed in Chinese pregnant women was CST I (41.9%), followed by CST IV-A (31.1%), CST III (18.7%), and CST IV-B (8.3%). Samples of pregnancy source distributed in all types of CSTs, but mainly in CST I and CST III. In contrast, postpartum samples were basically found in CST IV, especially had greater numbers of CST IV-A (*χ*^2^ = 92.08, *P* < 0.001). These results indicate that in Chinese women, vaginal microbiome lacks a portion of *Lactobacillus* after delivery, which makes the microbial diversity becoming higher, but it is not completely occupied by harmful bacteria such as *Gardnerella*.

Neither significant difference in the α and β diversity (weighted UniFrac, ANOSIM R = -0.009, *P* = 0.693) (Supplementary Figure S4A-F), nor bacterial taxa differed in relative abundance was observed between early and late stages of pregnancy (*P* > 0.05 after FDR correction, Supplementary Figure S4G-H).

### The vaginal microbiome during pregnancy varied by hypertensive disorders and abortion history

During pregnancy, the α-diversity of vaginal microbiome was significantly higher in pregnant women with hypertensive disorders (HBP) than without (*P* = 0.037, Figure 2A). The relative abundance of more than 30 bacterial taxa varied significantly between HBP and control groups based on linear discriminant analysis effect size (LEfSe) analysis (Figure 2B), and almost all of these discriminating taxa were enriched rather than depleted in the case group. Suffering from HBP was not associated with a reduction in the relative abundance of *Lactobacillus*, but was accompanied by an increase in proportion of genera such as *Gardnerella, Atopobium*, and *Sneathia*. The results reveal that HBP may have some impact on the vaginal microbiota during pregnancy, causing abundance changes in many bacteria, and part of which was consistent with that happened after delivery (Figure 1D).

**Figure 2.**
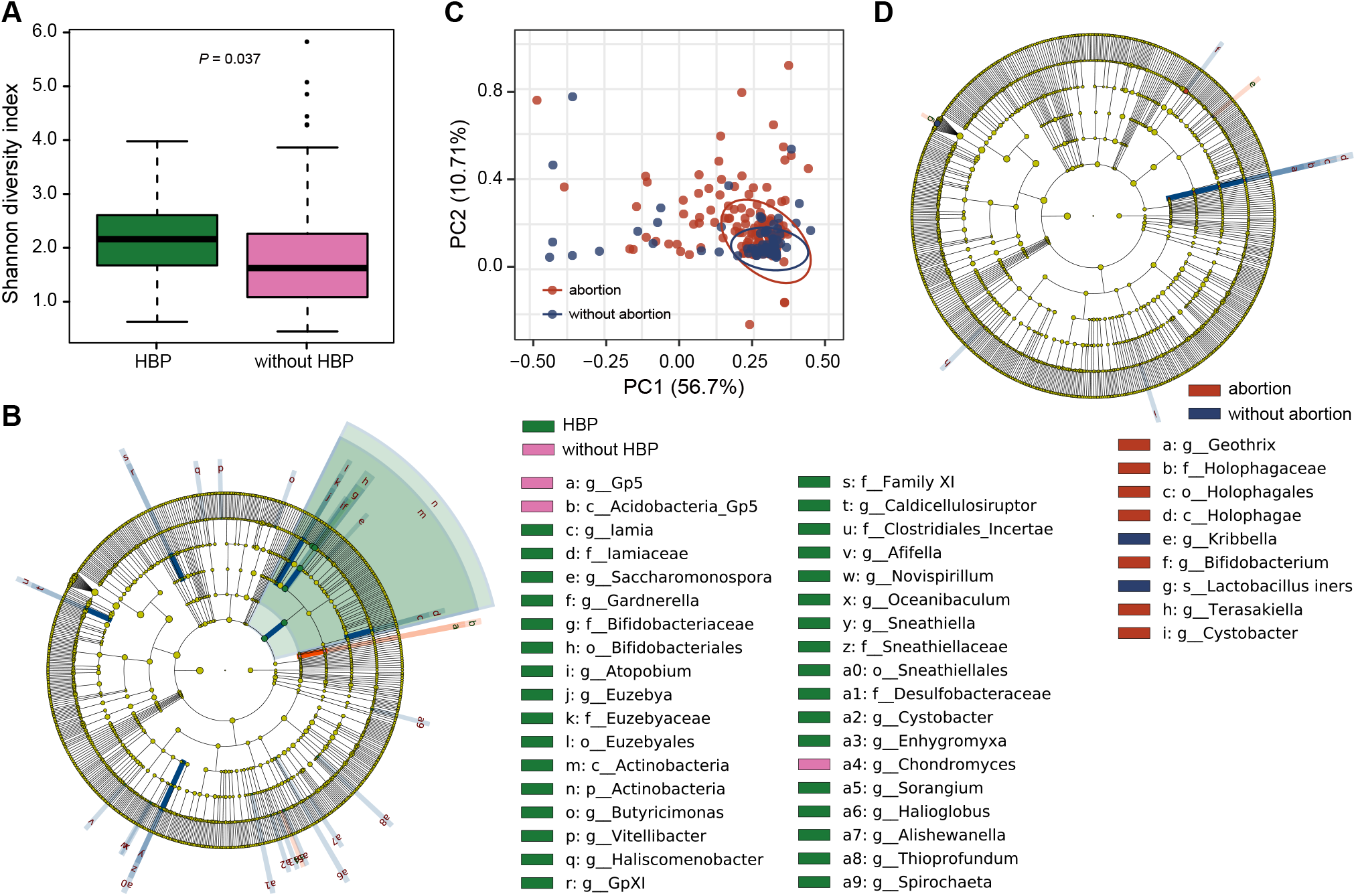
The vaginal microbiome of pregnant women with a history of abortion or hypertensive disorders changed during pregnancy. **(A)** Alpha diversity of the vaginal microbiome during pregnancy between pregnant women with (HBP) and without hypertensive disorders (without HBP). Each box plot represents the median, interquartile range, minimum, and maximum values. **(B)** Cladogram using LEfSe indicates the phylogenetic distribution of vaginal microbiome associated with pregnant women who had hypertensive disorders with pregnancy. **(C)** Weighted ANOSIMs based on the distance matrix of UniFrac dissimilarity of the vaginal microbiome during pregnancy in pregnant women with or without abortion history. The axes represent the two dimensions explaining the greatest proportion of variance in the communities. Each dot represents a sample, and each circle shows a 95% confidence interval. **(D)** Cladogram using LEfSe indicates the phylogenetic distribution of vaginal microbiome associated with pregnant women who had history of abortion. The LDA scores (log10) > 2 and *P* < 0.05 are listed. The prefixes p_, c_, o_, f_, g_, s_ represent phylum, class, order, family, genus, and species respectively.

Significant difference was found in community structure based on the weighted UniFrac (ANOSIM R = 0.054, *P* = 0.015) between pregnant women with and without history of abortion (Figure 2C). Samples collected from women who did not have abortion history were more intensive, while those with a history of abortion were discrete. When screening for the taxonomic distinction between the two groups, only 9 discriminating taxa were found (Figure 2D). The relative abundance of *L. iners* was significantly lower in pregnant women with abortion history. The clustering results indicate that the composition of vaginal microbiome is also related to abortion history. If the women had not experienced a surgical abortion in the past, their vaginal microbiota would be similar, on the contrary, abortion may increase the heterogeneities between individuals, resulting in completely different community structure. Because there were more discriminating taxa that have undergone significant changes, the impress of HBP on vaginal microbiome seems to be greater than the history of abortion. This may be partly due to the fact that HBP occurring during pregnancy has a greater impact on maternal physiology which would immediately transfer to the microbiota, while the previous abortion leaves only a slight trace.

Although there was no significant difference in α and β diversity, several characteristic bacteria corresponding to four factors including delivery mode, maternal age, gestational diabetes mellitus, and hypothyroidism could be found in the vaginal microbiome during pregnancy (Supplementary Figure S5).

### The vaginal microbiome postpartum varied by delivery mode and maternal age

Postpartum vaginal microbiome showed a strong association with delivery mode. After delivery, the microbial diversity of vaginal microbiome in the woman who delivered by cesarean section was significantly higher than vaginally delivered women (*P* < 0.001, Figure 3A). Significant difference was also found in community structure based on the weighted UniFrac between women who delivered cesarean section and vaginally, with the latter had a smaller β diversity (ANOSIM R = 0.080, *P* = 0.034, Figure 3B). It seems that the vaginal microbiota of the pregnant women who delivered vaginally is dominated by fewer microbial species after delivery, which also has a more similar community structure to each other. The discriminating taxa identified from the LEfSe analysis further reflect that the *Lactobacillus*, which are resident of healthy women’s vagina, were more abundant in the postpartum vagina of the pregnant women who delivered vaginally (Figure 3C).

**Figure 3.**
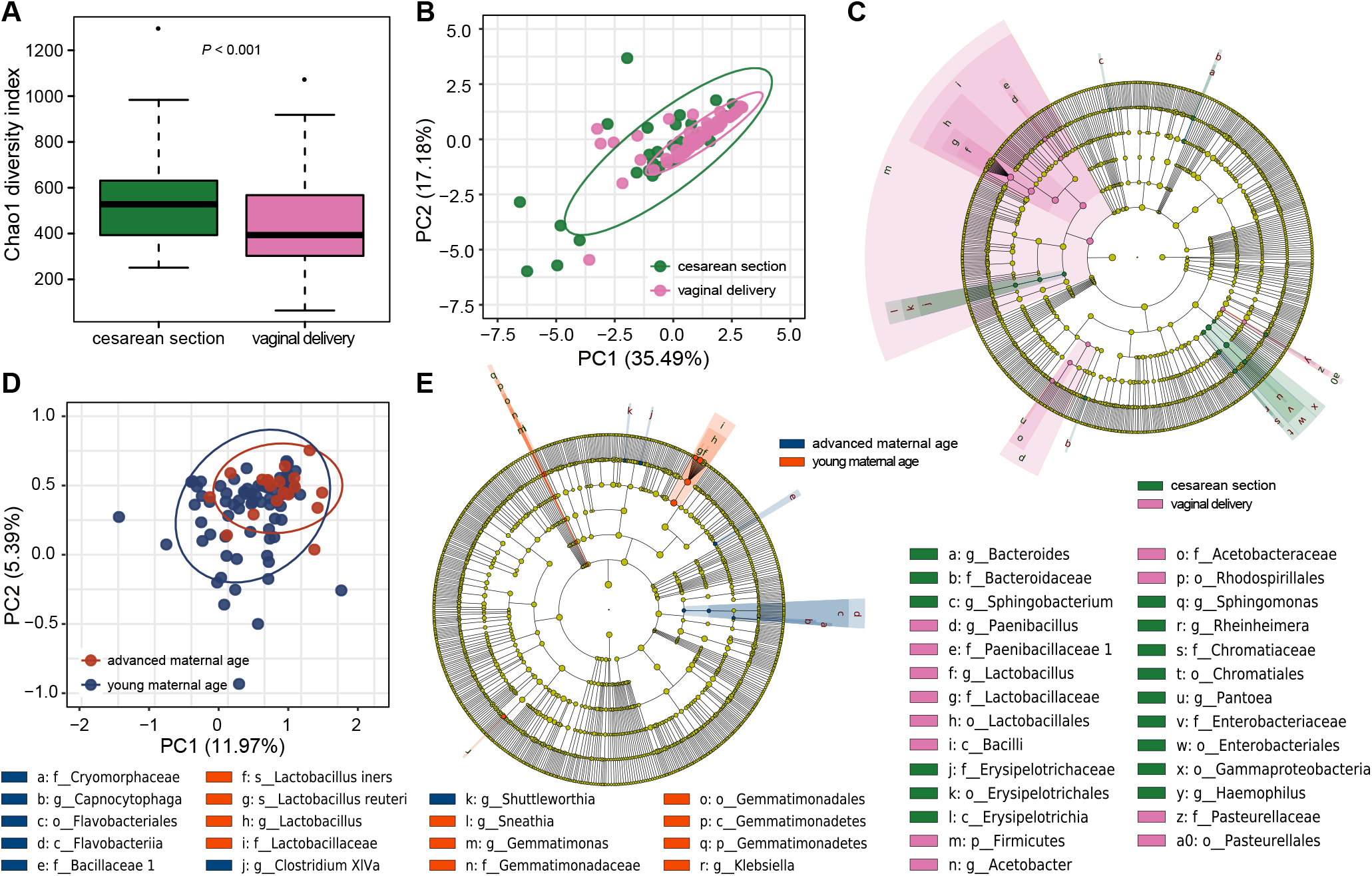
The vaginal microbiome postpartum varied by delivery mode and age of the pregnant woman. **(A)** Alpha diversity of the vaginal microbiome during pregnancy between the pregnant women who delivered naturally or cesarean section in the postpartum period. Each box plot represents the median, interquartile range, minimum, and maximum values. **(B)** Weighted ANOSIMs based on the distance matrix of UniFrac dissimilarity of the vaginal microbiome in the pregnant women who delivered naturally or cesarean section in the postpartum period. The axes represent the two dimensions explaining the greatest proportion of variance in the communities. Each dot represents a sample, and each circle shows a 95% confidence interval. **(C)** Cladogram using LEfSe indicates the phylogenetic distribution of vaginal microbiome associated with pregnant women who delivered naturally or cesarean section in the postpartum period. **(D)** Weighted ANOSIMs based on the distance matrix of UniFrac dissimilarity of the vaginal microbiome in the pregnant women who had advanced or young age in the postpartum period. **(E)** Cladogram using LEfSe indicates the phylogenetic distribution of vaginal microbiome associated with pregnant women who had advanced or young age in the postpartum period. The LDA scores (log10) > 2 and *P* < 0.05 are listed. The prefixes p_, c_, o_, f_, g_, s_ represent phylum, class, order, family, genus, and species respectively.

We found maternal age is another factor associated with the variation of postpartum vaginal microbiome. Samples collected from women with advanced and young age grouped into two distinct clusters (ANOSIM R = 0.149, *P* = 0.022, Figure 3C), and the elder formed a more consistent community structure. Based on LEfSe analysis, the vaginal microbiome of the women with advanced age were lacking *Lactobacillus* genus, and its two species *L. iners* and *L. reuteri* after delivery (Figure 3E). Besides delivery mode and maternal age, dramatic shifts in relative abundance of some bacteria were observed in the grouping of three other factors including abortion history, maternal pre-pregnancy weight status and pregnancy complications (Supplementary Figure S6).

### Identification of common key taxa accounting for the variation of vaginal microbiome

Totally, the relative abundances of 163 bacterial taxa from phylum to genus level varied significantly between groups during pregnancy and postpartum based on LEfSe analysis (Supplementary Figure S7). These bacterial taxa rarely had overlaps across different factors, and most of the discriminating taxa were unique to each abnormal factor. Moreover, the discriminating taxa of the same factor had no consistency between prenatal and postpartum. Certain bacterial taxa associated with maternal age even showed a completely opposite trend before and after delivery. The distinction of the discriminating taxa and the abundance divergence of the same taxa reflected the large differences of vaginal microbial communities during pregnancy and postpartum as well as their response to various abnormal factors.

Even though, several common taxa accounting for the variation of vaginal microbiome can still be identified from the 163 discriminating taxa. Relative abundance of 21 bacterial taxa was simultaneously altered in at least 2 of the 10 comparisons (Figure 4A). Thirteen bacterial taxa shifted in the same direction in two comparisons, and significant differences were consistently recorded for 8 genera *Propionibacterium, Rheinheimera, Butyricimonas, Lactobacillus, Sneathia, Bulleidia, Cellulosilyticum*, and *Nosocomiicoccus*. Common taxa were more likely to present with the two factors of delivery mode and maternal age, especially the postpartum microbiome associated with maternal age. In addition, Lactobacillaceae family decreased in abundance in pregnant women who delivered cesarean section and advanced maternal age (Figure 4B). The relative abundance of *L. iners* was lowered in pregnant women who delivered without abortion history during pregnancy and those with advanced maternal age in postpartum (Figure 4C). The abundances of *L. reuteri* depleted in women with abortion history and advanced maternal age in postpartum (Figure 4D). These results suggest that vaginal delivery, young maternal age, and without abortion history perhaps are more appropriate for the growth of *Lactobacillus* in maternal vagina, which should be good for women’s health. Some vaginal microbes showed the same response to different abnormal factors and may be more susceptible to the influence, so these bacteria together with the maternal age, require special care.

**Figure 4.**
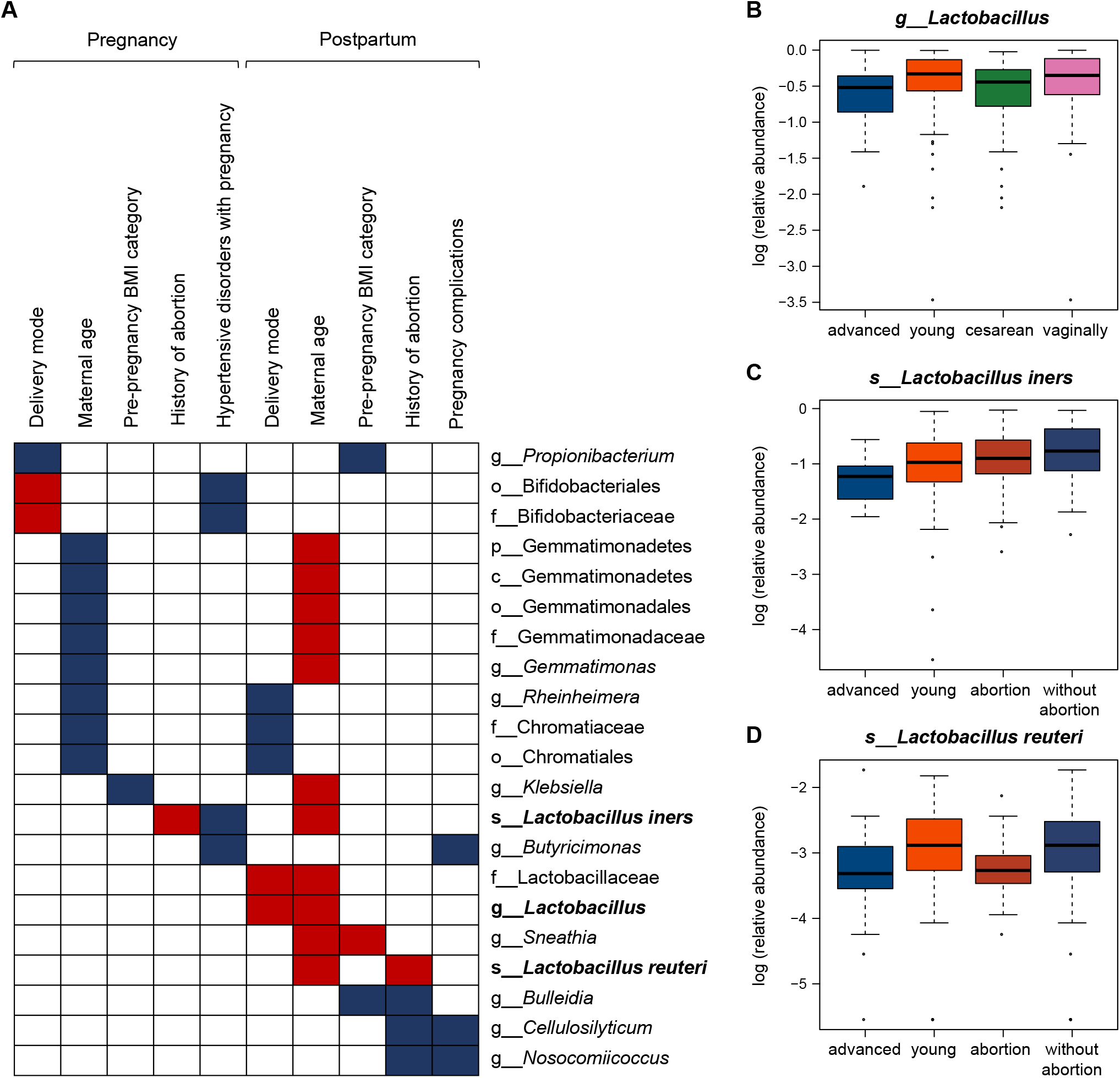
The abundance of specific vaginal bacteria changed in multiple groups during pregnancy and postpartum. **(A)** The relative abundances of the 21 bacterial taxa was varied significantly in two comparisons simultaneously during pregnancy and in postpartum period based on LEfSe analysis. The discriminating taxa were identified with a threshold of LDA scores (log10) > 2 and *P* < 0.05. The boxes filled in blue and red color represent the discriminating taxa enriched and depleted in the vaginal microbiome of women who delivered cesarean, with advanced maternal age, overweight, hypertensive disorders or other pregnancy complications, and without abortion history during pregnancy and postpartum, respectively. **(B)** The relative abundance of Lactobacillaceae family in pregnant women who delivered cesarean section and advanced maternal age. **(C)** The relative abundance of *Lactobacillus iners* in pregnant women without abortion history during pregnancy and those with advanced maternal age in postpartum. **(D)** The relative abundance of *Lactobacillus reuteri* in women without abortion history and young maternal age in postpartum. The prefixes p_, c_, o_, f_, g_, s_ represent phylum, class, order, family, genus, and species respectively.

### Variations of CSTs associated with multiple factors during pregnancy and postpartum

To facilitate the comparison of the relationships between vaginal microbiome and different factors in different periods, we once again examined the distribution of CSTs (Figure 5 and Supplementary Table S3). We found the largest discrimination before and after delivery was that CST I and CST IV-A had the highest proportions, respectively. In the vaginal microbiome during pregnancy, the ratios of CST IV-A in the groups of cesarean section, advanced maternal age, with abortion history, overweight, with hypertensive disorders, and with hypothyroidism were remarkably higher than those of corresponding groups. It is worth noting the difference of CSTs composition between pregnant women who have normal (term) delivery compared to PTB and PROM. The term delivery group during pregnancy showed a lower CST IV-A/CST I ratio (0.35), and such ratio increased to 0.53 if PROM occurred even though there was no premature birth at last (term-PROM). In addition, a high CST IV-A/CST I ratio (1.00) was shown in the group of pregnant women who have delivered PTB, and such ratio (2.00) was particularly high in PROM complicated with PTB (PROM-preterm) during pregnancy. In the vaginal microbiome after delivery, the situation of more CST IV-A was further exacerbated, with the only exception being hypertensive disorders. The divergence between term and PTB had not disappeared, and CST I was not even detected in both PROM-preterm and preterm.

**Figure 5.**
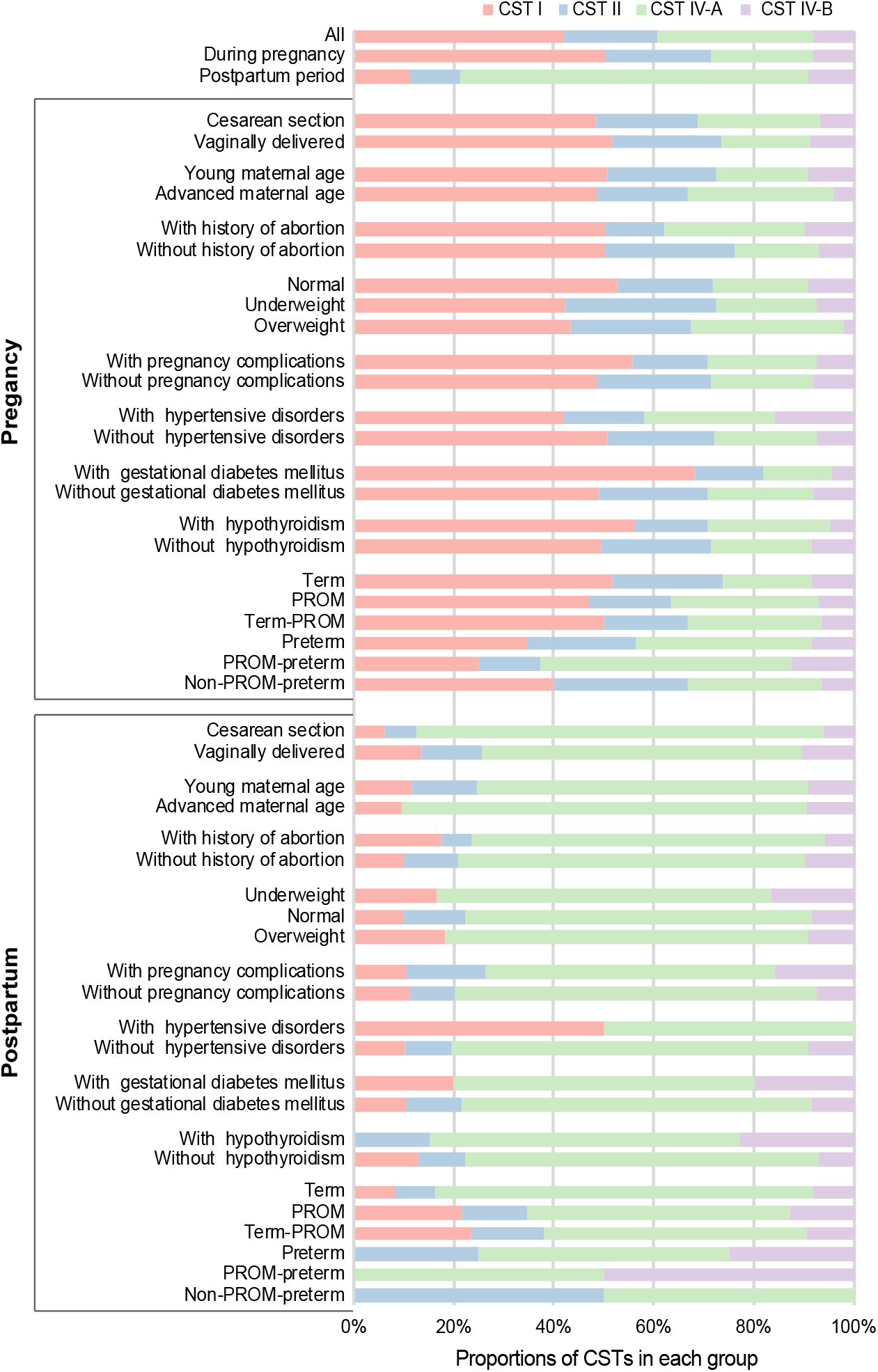
The prevalence of community state type (CST) in the vaginal microbiome was different corresponding to different maternal factors. The percentage of each CST in each group was represented by one color. The top three bars showed the mean of CST percentage in samples of all, during pregnancy, and postpartum period, respectively. Term denotes the pregnant woman who delivered in term, Term-PROM denotes premature rupture of membranes (PROM) occurred but birth in term at last, PROM-preterm denotes PROM complicated with preterm birth (PTB), and Non-PROM-preterm denotes without PROM occurred but premature birth at last.

To further investigate the connections between vaginal microbiome and PTB and PROM, microbial composition was compared between each group during pregnancy and postpartum. The relative abundances of 68 bacterial taxa, including 1 phylum, 3 classes, 9 orders, 18 families, 34 genera, and 3 species varied significantly between groups (Supplementary Figure S8A-G). During pregnancy, 6 bacterial taxa, including *Lactobacillus buchneri* and *Lactobacillus coryniformis*, and 2 bacterial taxa were increased in abundance in pregnant women with preterm or PROM neonate (Supplementary Figure S8A-B). When we stratified women who delivered preterm by PROM status, 3 bacterial taxa were associated with PROM related preterm (Supplementary Figure S8C), and 16 bacterial taxa were associated with non-PROM related preterm (Supplementary Figure S8D). Among women who delivered preterm, the abundances of 4 bacterial taxa were significantly higher in those with PROM compared to those without PROM (Supplementary Figure S8E). These discriminant bacteria in the vaginal microbiome, with significant changes in prenatal abundance, may be served as candidate biomarkers for predicting preterm birth. In postpartum, 20 bacterial taxa, such as *L. gasseri*, were more abundant in preterm group, while 9 bacterial taxa, such as *Bacillus*, were more abundant in term group (Supplementary Figure S8F). Significant differences were also recorded for the 19 bacterial taxa between pregnant women with and without PROM (Supplementary Figure S8G).

## Discussion

Both cross-sectional and longitudinal studies reported that the vaginal microbiome during pregnancy was less diverse and more stable than that of postpartum [20]. Chinese women have not been well represented before. To this end, this study characterized the vaginal microbiome of the largest cohort of Chinese women to date. Consistent with previous studies in women of African, Hispanic or European [10, 20], we also found that diversity and composition of vaginal microbial communities are relatively stable at early and late time points in pregnancy and dramatically changed in postpartum period to be less *Lactobacillus* dominant in Chinese population. Furthermore, we found that the vaginal microbiome changed in response to abortion history and hypertensive disorders during pregnancy and delivery mode and maternal age in the postpartum period.

Identification of key taxa within the diverse vaginal microbiome is of great importance because they may pose differing risks for adverse health outcomes in pregnant women. Our study identified a series of significant bacterial taxa that differed significantly in relative abundance according to the delivery mode, maternal age, history of abortion, and pregnancy complications. Notably, we also found several key taxa that differed significantly in relative abundance in at least two comparisons. Considering that these key taxa were constantly mentioned to be associated with women’s health in previous studies [6, 11], which point toward them as taxa of particular importance for study in the future. Other taxa, such as *Propionibacterium* enriched in cesarean section newborns [21], *Butyricimonas* enriched in the patients of Autism spectrum disorder [22] and depleted in patients with histamine intolerance [23] and thyroid cancer [24], and *Bulleidia* appears more frequently in patients with esophageal squamous cell carcinoma [25], also merit additional investigation.

CST is widely used in the studies of vaginal microbiome to deal with inter-subject and/or intra-subject variability of women [26]. Through a comparison of this new dataset with existing datasets from Caucasian women and other countries, it revealed that the prevalence of CSTs may be different across populations. Using a large sample size of 396 North American women from four ethnic groups, including white, black, Hispanic, and Asian, Ravel et al. established five CSTs [18]. Huang et al reported the identification of four CSTs from 34 Chinese women during different pregnancy stages [27]. Considering that there is a distinct lack of information regarding the CSTs in Chinese pregnant women, our results from a population of large Chinese women contribute to a more comprehensive understanding of CSTs of Chinese pregnant women. Similar to Huang et al, we also identified four CSTs previously described: I (41.9%), III (18.7%), IV-A (31.1%), and CST IV-B (8.3%), but did not find CST II and CST V in our study population. The possible reasons of lack some CSTs in Chinese women and their difference to Caucasian women may be their living environment or ethnicity.

Unexpectedly, we found that changes in the vaginal microbiome community is also related to GDM, which has not been reported in neither Chinese nor Caucasian population. Compared with healthy pregnant women, we found that women suffered from GDM had a higher proportion of CST I during pregnancy, while the proportion of non-*Lactobacillus* dominated CST IV was lower. After delivery, the proportion of CST I in GDM women was still higher than that of healthy women. But unlike during pregnancy, the proportion of CST IV-B which contains more *Gardnerella* and is generally considered to be unfavorable to women’s vaginal health [18], increased remarkably in postpartum women who have had GDM. These results suggest that GDM may be more harmful to postpartum vaginal microbiome and health than to prenatal, and more attentions should be paid to the postpartum health of this female population.

In this study, we present a study design with the challenge of mixed and even discordant findings in some studies linking the vaginal microbiome to PTB in mind. Consistent with the Romero’s study [4], we found no differences in the frequency of observed CSTs between women who delivered at term and those who delivered preterm. However, compared to their results which did not find key taxa differed in relative abundance [4], we found bacterial taxa were significantly different between PROM related PTB and non-PROM related PTB, indicating that different types of PTB had specific bacterial taxa. Some of contributing taxa such as *Gardnerella, Ureaplasma* and/or *Megasphaera* could be considered for developing predictive models in Chinese population. We can replicate previously reported association between more *Gardnerella, Ureaplasma* or *Megasphaera* and clinically heterogeneous PTB in cohorts of predominantly African descent in postpartum stage [5, 7]. The previously hypothesized associations between less *Lactobacillus* abundance and PTB in cohorts of predominantly African descent, more *L. iners* abundance and PTB in cohort of predominantly European descent were not replicated in our study [5-7]. The differences between previous studies and our results strongly suggest that PBT-microbiome associations may be racially-dependent. Further population-specific studies are needed to assess the impacts of the association between vaginal microbiome and PTB and to identify population-specific key taxa.

Although the present study represents the most extensive examination of the vaginal microbiome of Chinese pregnant women to date, in reviewing of our results, several limitations should be considered. First, this is a cross-sectional study in which samples were obtained at a single time point. Looking forward, prospective longitudinal studies are also needed to confirmed our results. Secondly, studies using metagenomic sequencing are needed in the future to provide more detailed information about the function and changes in the vaginal microbiome. In addition, absence of a concurrent analysis of host factors is a likely reason for the variable conclusions [28].

## Conclusions

This is the first study to characterize a Chinese cohort of this size. We identified measurable differences in vaginal microbiome of Chinese pregnant women according to delivery mode, maternal age, and history of abortion and hypertensive disorders, with possible consequences for both short- and long-term health. The study also demonstrated that PBT-microbiome associations are population-dependent and reveals new insights into ethnic and biogeographical effects upon the association between PTB and vaginal microbiome. With the realization that our understanding of the microbiome is as good as the diversity of people sampled, this dataset will provide valuable information for future research and contribute to a more comprehensive understanding of the correlation between bacterial community and Chinese pregnant women.

## Materials and methods

### Patients and samples

The study was performed with the approval of the Ethical Committee of Beijing Institute of Microbiology and Epidemiology and conducted according to the principles expressed in the Declaration of Helsinki. From July to December 2016, totally 474 pregnant Chinese women attending the Department of Obstetrics at the 301 Hospital (Beijing) for regular check-ups were enrolled in this study. All participants provided informed consent. Information regarding demographic characteristics, medical history, clinical manifestation, laboratory test results were prospectively collected using a standard questionnaire. For each participant, a cotton swab was used to collect discharge from posterior vagina. The samples were stored at -20°C upon collection. All samples were stored at -80°C within 4 hours until metagenomic DNA extraction.

### Processing of microbial samples

Microbial DNA was extracted from vaginal swabs and sterile water using the DNA extraction kit (Cat No. 69504, Qiagen, Hilden, Germany) according to manufacturer’s protocols. DNA concentration and purity were measured by Qubit 3.0 (Cat No. Q33216, Thermo Fisher Scientific, Waltham, MA). DNA was amplified using polymerase chain reaction (PCR) with a pair of barcoded primers (341F: CCTAYGGGRBGCASCAG; 806R: GGACTACNNGGGTATCTAAT) targeting the V3-V4 region of 16S rRNA gene. Each PCR reaction was conducted in a 30µL reaction system with 15µL of Phusion High-Fidelity PCR Master Mix (Cat No. F531L, New England BioLabs, Ipswich, MA), 0.2µM of forward and reverse primers, and about 10ng DNA templates. Negative extraction controls and blank controls with sterile water as the PCR template were included. Same volume of 1X loading buffer (contained SYBR green) was mixed with PCR products and then electrophoresis was operated on 2% agarose gel for detection. Samples with bright main strip between 400-450bp were chosen for further experiments. PCR products were pooled in equimolar ratios. Then, the mixture was purified with GeneJET Gel Extraction Kit (Cat No. K0691, Thermo Fisher Scientific, Waltham, MA). Sequencing libraries were constructed using NEB Next Ultra DNA Library Prep Kit for Illumina (Cat No. E7370L, New England BioLabs, Ipswich, MA) following manufacturer’s recommendations. The library quality was assessed on the Agilent Bioanalyzer 2100 system (Agilent Technologies, Santa Clara, CA). At last, the library was sequenced on an Illumina HiSeq2500 platform (Illumina, San Diego, CA) and 250bp paired-end reads were generated.

### Bioinformatics and statistical analysis

Paired-end reads were merged into long sequences based on the overlaps between reads1 and reads2 by using FLASH [29]. Merged sequences then were analyzed using QIIME 1.9.1 software package [30]. First, sequences were filtered by QIIME quality filters. Then we used pick_de_novo_otus.py to pick OTUs in addition to generate an OTU table. Sequences with a ≥ 97% similarity were assigned to the same OTUs. A representative sequence was picked for each OTU and the RDP database was used to generate taxonomic information for each representative sequence.

To compute Alpha diversity, the OTU table was rarified and five metrics including Chao1, observed species, PD whole tree (Faith’s Phylogenetic Diversity, which adds up all the branch lengths of the phylogenetic tree as a measure of diversity), Shannon and Simpson were calculated. Rarefaction curves were generated based on these metrics. Both weighted and unweighted unifrac distances were calculated for principal coordinate analysis (PCoA). The pairwise dissimilarity between the microbial community structures was assessed using Bray-Curtis distance at the OTU level as described before [31]. The difference in microbial markers was measured using Mann-Whitney rank test and LEfSe. When multiple hypothesis tests were performed simultaneously, *P* values were corrected using Benjamini and Hochberg’s false discovery rate (FDR). For the comparative analysis, only the genera and species with the abundance of > 1 and > 0.2%, respectively, in at least one of the samples were included.

The clustering of CSTs was done using complete linkage hierarchical clustering with five clusters as described by Ravel et al [18, 20]. CST I, CST II, CST III and CST V was predominated with *L. crispatus, L. gasseri, L. iners*, and *L. jensenii*, respectively. CST IV was defined as lacking *Lactobacillus spp*. and comprising a diverse set of strict and facultative anaerobes, and was further split into CST IV-A and CST IV-B.

## Data Availability

The sequencing data has been submitted to GSA at the National Genomics Data Center, with the accession ID CRA002692. All data referred to in the manuscript would be made available upon request.

## Ethical statements

The study was performed with the approval of the Ethical Committee of Beijing Institute of Microbiology and Epidemiology and conducted according to the principles expressed in the Declaration of Helsinki. All participants provided informed consent.

## Data availability

The sequencing data has been submitted to GSA at the National Genomics Data Center, with the accession ID CRA002692.

## Authors’ contributions

FZ and WL conceived the study. XZ and JW designed the study and interpreted the data. XZ and JW wrote the paper. QZ and ZG collected the samples and conducted the experiments. XZ, JW, XM, BX, HF, FZ and WL analyzed the data and created the graphs. All authors approved the final version of the manuscript.

## Acknowledgments

This work was supported by the National Natural Science Foundation of China (81825019, 31722031, 31670119, 31870107), the Beijing Leading Talents in Science and Technology (Z181100006318008), the China Mega-Project on Infectious Disease Prevention (2018ZX10713002-002, 2018ZX10101003-002, 2018ZX10301401), and the National Key Research and Development Program of China (2016YFC1000705). We are grateful to all the subjects, their families, and collaborating clinicians for their participation.

## Supplementary material

**Table S1. Characteristics of pregnant women included in the study (DOCX 16 kb)**

**Table S2. The significantly different bacteria between pregnancy and postpartum (XLSX 20 kb)**

**Table S3. Distribution of community state types according to delivery mode, age, abortion, BMI, pregnancy and delivery complications (DOCX 25 kb)**

**Figure S1. The** α **diversities of the vaginal microbiome during pregnancy were significantly lower than those of postpartum**. Differences in the α diversities of the vaginal microbiome between pregnancy and postpartum according to the observed species, PD whole tree (Faith’s Phylogenetic Diversity, which adds up all the branch lengths of the phylogenetic tree as a measure of diversity), Shannon and Simpson index. Each box plot represents the median, interquartile range, minimum, and maximum values.

**Figure S2. The vaginal microbiome of pregnant women was more similar to each other during pregnancy**. Weighted and unweighted PCoA (A-B) and ANOSIM (C-D) based on the distance matrix of UniFrac dissimilarity of the vaginal microbial communities in pregnancy and postpartum. Respective ANOSIM R values show the community variation between the compared groups, and significant *P* values are indicated. The axes represent the two dimensions explaining the greatest proportion of variance in the communities. Each dot represents a sample.

**Figure S3. The vaginal microbiome of most women after delivery was classified as CST IV-A**. Color bar from yellow to red shows the relative abundance of each microbial taxon. The prefixes g_, s_ represent genus, and species respectively.

**Figure S4. Little difference was shown between early and late stages of pregnancy in microbial diversity, community structure and composition of the vaginal microbiome**. (A-E) Differences in the microbial diversities of the vaginal microbiome between early and late stages according to the Chao 1, observed species, PD whole tree, Shannon and Simpson index. (F) Weighted ANOSIMs and PCoA based on the distance matrix of UniFrac dissimilarity of the vaginal microbiome of early and late pregnant stages. (G) Relative abundance of the discriminating taxa in the vaginal microbiome between early and late pregnant stages at all levels. (H) Relative abundance of the discriminating taxa in the vaginal microbiome of early and late pregnant stages at the genus level. The discriminating taxa were identified based on LEfSe analysis with a threshold of LDA scores (log10) > 2 and *P* < 0.05. The prefixes p_, c_, o_, f_, g_, s_ represent phylum, class, order, family, genus, and species respectively.

**Figure S5. LEfSe analysis showed the discriminant bacteria of vaginal microbiome during pregnancy corresponding to multiple complex factors**. (A) Different delivery mode. (B) Advanced maternal age vs. young maternal age. (C) With vs. without gestational diabetes mellitus. (D) With vs. without hypothyroidism. The LDA scores (log10) > 2 and *P* < 0.05 are listed. The prefixes p_, c_, o_, f_, g_, s_ represent phylum, class, order, family, genus, and species respectively.

**Figure S6. LEfSe analysis showed the discriminant bacteria of vaginal microbiome in postpartum corresponding to multiple complex factors**. (A) Different body mass index before pregnancy. (B) With vs. without abortion history. (C) With vs. without pregnancy complications. The LDA scores (log10) > 2 and *P* < 0.05 are listed. The prefixes c_, o_, f_, g_, s_ represent class, order, family, genus, and species respectively.

**Figure S7. Most discriminant bacteria of the vaginal microbiome were different under complex factors during pregnancy and postpartum**. The relative abundance of 163 bacterial taxa, including 5 phyla, 12 classes, 22 orders, 35 families, 86 genera, and 3 species, varied significantly between groups during pregnancy and postpartum based on LEfSe analysis. The LDA scores (log10) > 2 and *P* < 0.05 are listed. The boxes filled in blue and red color represent the discriminating taxa enriched and depleted in the vaginal microbiome of women who delivered cesarean, with advanced maternal age, overweight, hypertensive disorders, hypothyroidism or other pregnancy complications, and without abortion history or gestational diabetes mellitus during pregnancy and postpartum, respectively. The prefixes p_, c_, o_, f_, g_, s_ represent phylum, class, order, family, genus, and species respectively.

**Figure S8. PTB and PROM were accompanied by some bacteria enrichment in the vaginal microbiome**. Based on LEfSe analysis, the relative abundances of 68 bacterial taxa varied significantly between groups. The LDA scores (log10) > 2 and *P* < 0.05 are listed. (A) Preterm vs. term group during pregnancy. (B) PROM vs. term group during pregnancy. (C) PROM related preterm vs. term group during pregnancy. (D) Non-PROM related preterm vs. term group during pregnancy. (E) PROM related preterm vs. non-PROM related preterm during pregnancy. (F) Preterm vs. term group in postpartum. (G) PROM vs. term group in postpartum. The prefixes p_, c_, o_, f_, g_, s_ represent phylum, class, order, family, genus, and species respectively.

